# Examining Recent Trends in the Racial Disparity Gap in Tap Water Consumption: NHANES 2011–2018

**DOI:** 10.1101/2021.04.06.21255016

**Authors:** Asher Y. Rosinger, Anisha I. Patel, Francesca Weaks

## Abstract

**Objective:** As tap water distrust has grown in the US with greater levels among Black and Hispanic households, we aimed to examine recent trends in not drinking tap water including the period covering the US Flint Water Crisis and racial/ethnic disparities in these trends.

**Design, Setting, and Participants:** We analyzed data from the cross-sectional National Health and Nutrition Examination Survey data (2011–2018) for 9,439 children aged 2-19 and 17,268 adults. Log-binomial regressions and marginal predicted probabilities examined US nationally-representative trends in tap and bottled water consumption overall and by race/ethnicity.

**Results:** Among US children and adults, respectively, in 2017-2018 there was a 63% (adjusted prevalence ratio [PR]:1.63, 95%CI: 1.25-2.12, p<0.001) and 40% (PR:1.40, 95%CI: 1.16-1.69, p=0.001) higher prevalence of not drinking tap water compared to 2013-2014 (pre-Flint Water Crisis). For Black children and adults, the probability of not drinking tap water increased significantly from 18.1% (95%CI: 13.4-22.8) and 24.6% (95%CI: 20.7-28.4) in 2013–14 to 29.3% (95%CI: 23.5-35.1) and 34.5% (95%CI: 29.4-39.6) in 2017–2018. Among Hispanic children and adults, not drinking tap water increased significantly from 24.5% (95%CI: 19.4-29.6) and 27.1% (95%CI: 23.0-31.2) in 2013-14 to 39.7% (95%CI: 32.7-46.8) and 38.1% (95%CI: 33.0-43.1) in 2017-2018. No significant increases were observed among Asian or white persons between 2013-14 and 2017-18. Similar trends were found in bottled water consumption.

**Conclusions:** This study found persistent disparities in the tap water consumption gap from 2011–2018. Black and Hispanics’ probability of not drinking tap water increased following the Flint Water Crisis.

## Introduction

Approximately two million Americans lack basic access to drinking water^1^. However, this figure does not capture individuals who have access but do not drink their tap water. Not drinking tap water occurs for many reasons, including aesthetic considerations or distrust due to water quality violations in homes^2^ or schools^3^. The Flint, Michigan Water Crisis^4^, in which the Flint community was exposed to high concentrations of lead in their water from 2015 through 2016 raised national awareness^5^ of tap water safety concerns in US communities^2,6^. Then, Newark, New Jersey reported a lead crisis in 2016^7^ further stoking fears. Both Flint and Newark are comprised predominantly of non-white residents.

Other households in the US lack consistent access to tap water at home because of water shutoffs due to affordability issues^8^, which disproportionately affect residents in low-income housing, and Black and Hispanic households^9,10^. The differential treatment of individuals in these categories increases exposure to environmental injustice and racism^11^ which may affect both access to and use of tap water. Consequently, these groups are also more likely to drink bottled water^12^ and sugar-sweetened beverages^13,14^ that are more costly^15^ and lack the protective benefits of fluoride^16^ in tap water.

While previous work documented an increase in US children not drinking tap water in 2015-2016^12^ during the Flint Water Crisis compared to prior years, it is critical to surveil this nutritional behavior as a way to monitor trust and usage of tap water. Therefore, we examined whether not drinking tap water in the US continued to increase in 2017-2018 by analyzing nationally-representative trends among children/adolescents and adults, and whether disparities in this behavior persisted by race/ethnicity.

## Methods

Data come from the National Health and Nutrition Examination Survey (NHANES)^17^, a cross-sectional survey of the civilian, non-institutionalized household population, which uses a complex, multistage probability design. Detailed sampling procedures and methodology are described elsewhere^18^. NHANES is conducted by the National Center for Health Statistics (NCHS) and approved by their research ethics board. Children aged 7-17 years provided assent and parents provided consent for children under 18; adults consented for themselves.

This analysis uses the four most recent two-year cycles (2011–2018) because oversampling of non-Hispanic (NH) Asians began in 2011-12 allowing for an examination of this population’s trends in tap water consumption, and because prior work already examined trends in Black, Hispanic and white tap water consumption from 2007-2016^12^. Response rates ranged from 77.0% in 2011–12 to 54.6% in 2017–18 for the examination component for youth aged 1-19 years, and from 64.5% in 2011–12 to 45.3% in 2017–18 for adults aged 20+.

To assess the prevalence of not drinking tap water, we analyzed responses to the dietary recall question: “When you drink tap water, what is the main source of the tap water?” If respondents reported not drinking tap water, they were coded as “did not drink tap.” All others were coded as “drank tap water”^16,19^. A proxy responded to interview questions for children aged ≤5 years; interviews for children aged 6–11 years were conducted with proxy assistance while children aged ≥12 years responded themselves. If they responded that they did not know their primary source, they were excluded from analyses.

For robustness analyses, we analyzed day-one 24-hour dietary recall data and dichotomized tap water consumption and bottled water consumption as >0 ml or 0 ml following prior work^13^. This allowed for a comparison of consumption on a given day to self-reports of not drinking water from their main source of tap and if not drinking tap water shifted consumption to bottled water.

### Statistical analyses

A 2-sided *P*-value of .05 was used to assess statistical significance. Multiple log-binomial regression models were estimated since prevalence of not drinking tap was >10%^12^. As there were no significant race/ethnicity by survey cycle interactions (all p>0.05), we estimated the regressions and covariate-adjusted time-trends without interaction terms. We used the 2013-14 survey cycle as the reference category as it was the last cycle prior to the Flint Water Crisis. We plotted trends from the log-binomial regressions using marginal standardization^20^ to generate predicted probabilities by self-reported race/ethnicity (white, Black, Asian, Hispanic, or other). We adjusted for the range^21^ of the following covariates: age (2-5, 6-11, 12-19, 20-39, 40-59, 60+), sex, federal income to poverty ratio (FIPR; ratio of family income to the federal poverty guideline^22^: ≤130%, 131-350%, >350%), household reference educational attainment (for children/adolescents; high school graduate/GED equivalent and some college were released combined) and educational attainment (for adults), and whether respondents were born in the US or not^12,19^.

As robustness analyses, we re-estimated log-binomial regressions models as described above with the dichotomized outcome of not drinking any tap water (0 ml) on a given day using the 24-hour dietary recall. We repeated this analysis with the dichotomized outcome of any (>0 ml) bottled water intake.

All analyses accounted for the complex survey design, including day-one dietary sample weights, which adjusted for oversampling, non-response, non-coverage, and day of week^18,23^ and incorporated guidelines from NCHS^24^. Statistical analyses were conducted using Stata V15.1 (Statacorp, College Station, TX).

## Results

Data for the primary analyses were analyzed for 9,439 children and 17,268 adults with complete covariate information (**Table 1**). Overall, the probability of not drinking tap increased significantly from 12.3% (95% confidence interval [CI]: 9.5-15.0) and 13.9% (95% CI: 11.8-16.0) among children and adults, respectively in 2013-2014 to 19.9% (95% CI: 16.0-23.8) and 19.5% (95% CI: 16.5-22.5) in 2017-2018. Among children/adolescents and adults, respectively, in the post-Flint period (2017-2018) there was a 63% (adjusted prevalence ratio [PR]: 1.63, 95% CI: 1.25-2.12, p<0.001) and 40% (PR: 1.40, 95% CI: 1.16-1.69, p=0.001) higher prevalence of not drinking tap water compared to pre-Flint (2013-2014), respectively (**Figure 1a-b; Supplemental Table 1**: Models 1-2).

**Table 1:** Descriptive characteristics of US children/adolescents and adults, NHANES 2011-2018

|  | Children/adolescents <sup>a,c</sup> (n=9,439) | Adults <sup>a,c</sup> (n=17,268) |
| --- | --- | --- |
|  | <sup>b</sup>Mean | <sup>b</sup>Mean |
| 2011-12 | 25.9% | 24.4% |
| 2013-14 | 25.3% | 25.2% |
| 2015-16 | 25.0% | 25.2% |
| 2017-18 | 23.8% | 25.2% |
| % Don't drink tap from main water source | 15.0% | 16.2% |
| % Didn't drink tap on a given day | 48.6% | 46.3% |
| % Drink bottled water on a given day | 37.1% | 37.5% |
| % Male | 48.5% | 51.4% |
| 2-5 years / 20-39 years | 22.3% | 35.9% |
| 6-11 years / 40-59 years | 35.6% | 36.8% |
| 12-19 years / 60+ years | 42.1% | 27.3% |
| NH white | 54.7% | 66.6% |
| NH Black | 13.5% | 10.8% |
| Asian | 4.0% | 5.2% |
| Hispanic | 22.0% | 13.9% |
| % Born in US | 95.2% | 83.9% |
| FIPR: ≤130% | 34.9% | 22.6% |
| 131-350% | 37.3% | 35.2% |
| >350% | 27.7% | 42.2% |
| Adult education: | -- |  |
| Less than high school |  | 13.0% |
| High school grad | -- | 22.7% |
| Some college | -- | 32.8% |
| College + | -- | 31.4% |
| HH Ref education: |  | -- |
| Less than high school | 17.8% |  |
| High school grad/some college | 54.3% | -- |
| College + | 27.9% | -- |

<sup>a</sup>Unweighted sample size. <sup>b</sup>Weighted mean percentages for each category. <sup>c</sup>Without missing covariate data and valid dietary recall status. Other race/Hispanic origin included in analyses but not shown. FIPR: Federal income poverty ratio

**Figure 1:**
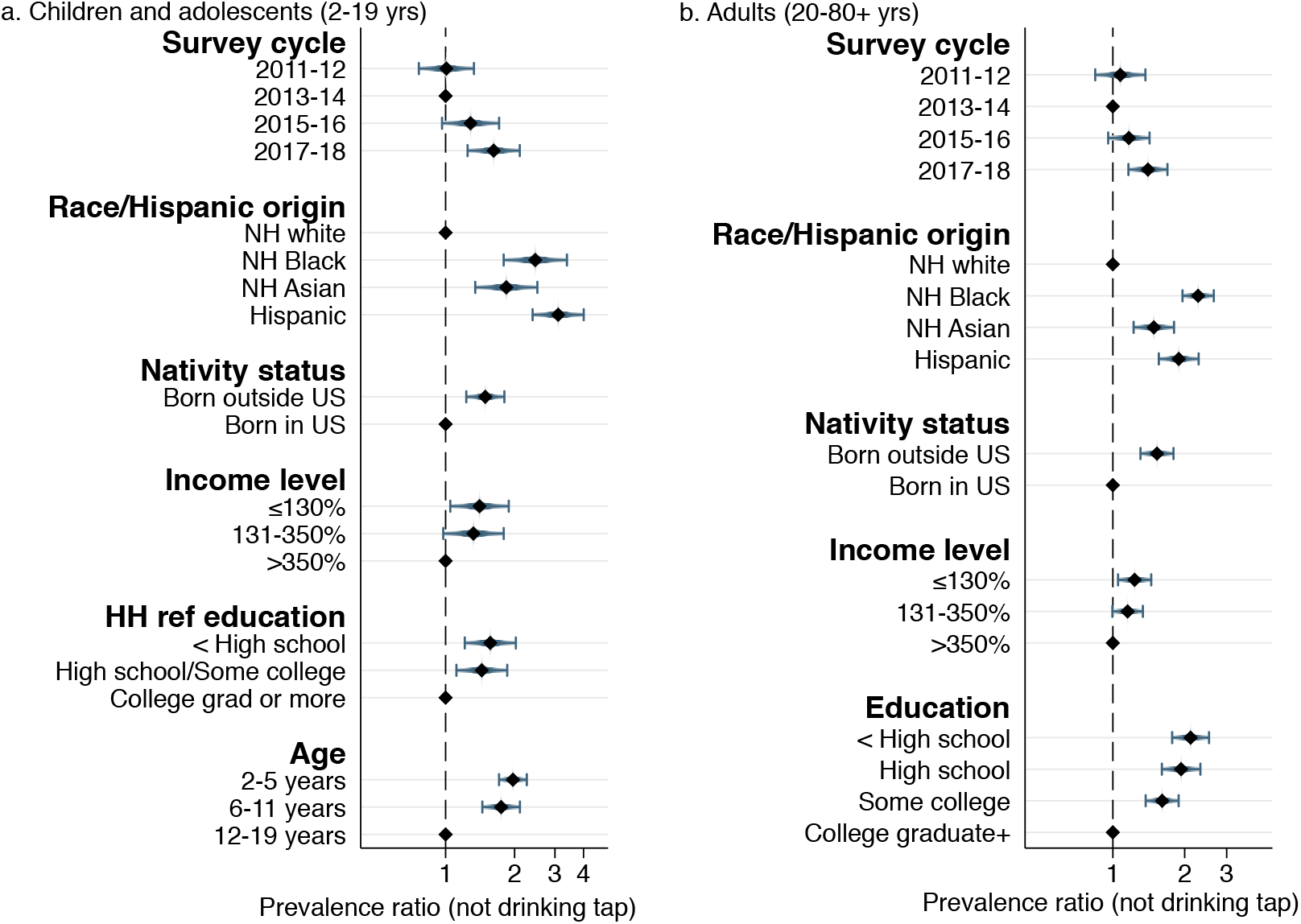
Log-binomial regression models of not drinking tap water by survey cycle, race/ethnicity, and socio-demographics, NHANES 2011-2018 among (a) children/adolescents and (b) adults. **Notes:** n=9,439 children/adolescents aged 2-19; n=17,268 adults; models adjusted for all variables shown in addition to sex, and age (for adults). Full models shown in Supplemental Table 1.

Hispanic, Black, and Asian children/adolescents and adults had significantly higher prevalence of not drinking tap than white persons (all p<0.001, **Figure 1a-1b; Supplemental Table 1:** Models 1-2). Lower income, less educational attainment, and being born outside the US were associated with higher prevalence of not drinking tap water (**Figure 1a-1b**).

Covariate-adjusted trends by race/ethnicity of not drinking tap water are presented in **Figure 2a-2b**. Among Black children/adolescents and adults, the probability of not drinking tap water increased significantly from 18.1% (95% CI: 13.4-22.8) and 24.6% (95% CI: 20.7-28.4) in 2013–14 to 29.3% (95% CI: 23.5-35.1) and 34.5% (95% CI :29.4-39.6) in 2017-18, respectively.

**Figure 2:**
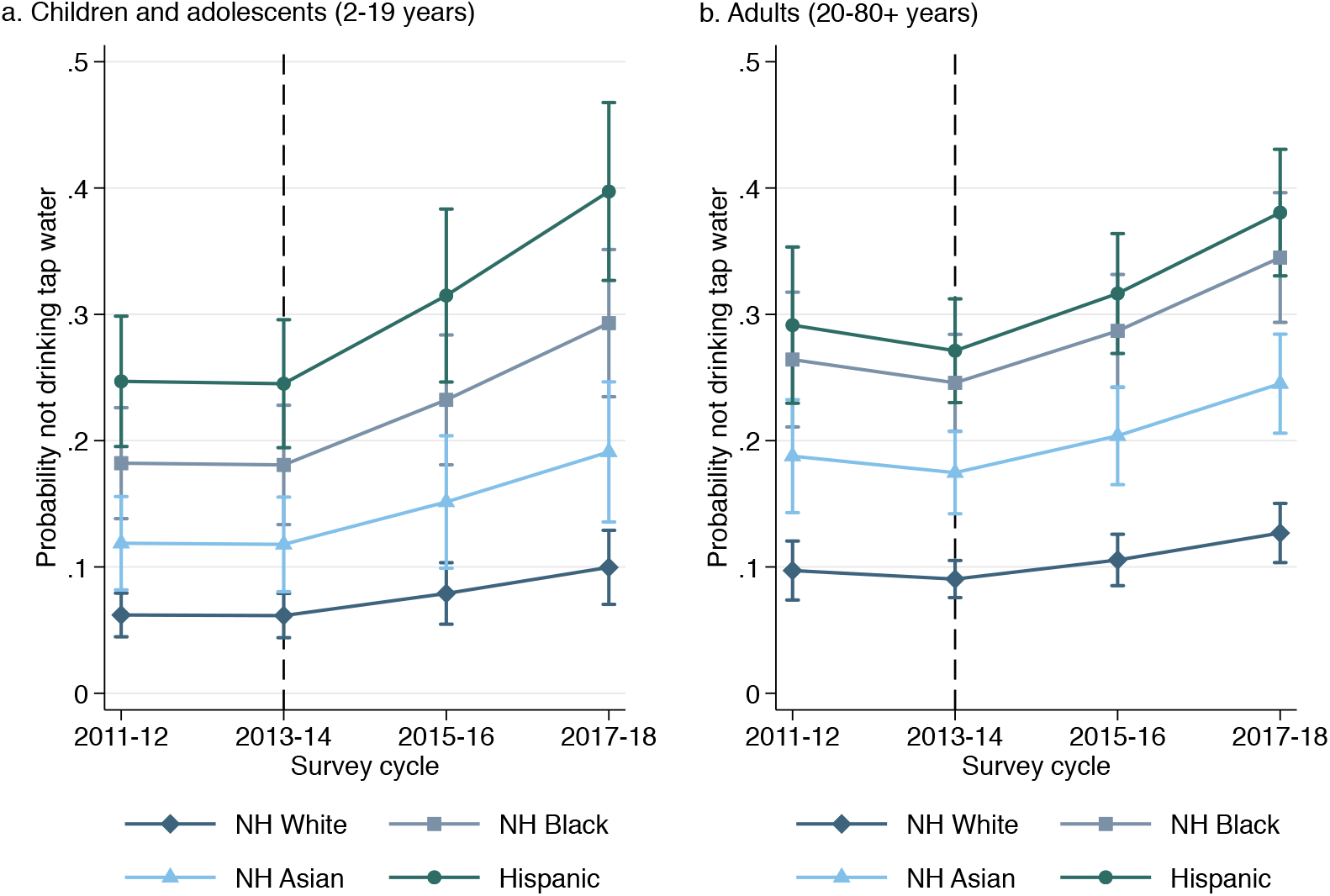
Covariate-adjusted predicted probability time trends and 95% confidence intervals of not drinking tap water by race/ethnicity among (a) children and (b) adults, NHANES 2011-2018. **Notes:** n=9,439 children aged 2-19; n=17,268 adults; models adjusted for nativity status, sex, age group, income level (federal income to poverty ratio), and educational attainment (of household reference for children, self for adults); dashed line at 2013–2014 indicates beginning of Flint, Michigan Water Crisis. Built from models shown in Supplemental Table 1.

Among Hispanic children/adolescents and adults, not drinking tap increased from 24.5% (95% CI: 19.4-29.6) and 27.1% (95% CI: 23.0-31.2) in 2013-14 to 39.7% (95% CI: 32.7-46.8) and 38.1% (95% CI: 33.0-43.1) in 2017–2018, respectively (**Figure 2a-2b**). The probability of not drinking tap water did not significantly increase during between 2013-14 and 2017-18 among Asian or non-Hispanic white children/adolescents and adults, respectively.

### Robustness analysis

In robustness analyses, we used data from 10,254 children/adolescents and 17,829 adults with valid dietary recall data, slightly more than the primary analyses due to fewer missing responses. Results of the log-binomial regressions examining not drinking tap water on a given day were slightly attenuated but largely consistent with main analyses (**Table 3**). Overall for children/adolescents and adults, the prevalence of not drinking tap water was 14.0% (PR=1.14; 95% CI: 0.98-1.33) and 12.6% (PR=1.13; 95% CI: 1.01-1.25) higher in 2017-2018 than 2013-2014. As in primary analyses, Black and Hispanic children/adolescents and adults had significantly higher prevalence of not drinking any tap water on a given day than white persons (**Table 3**, Models 1-2).

**Table 2:**
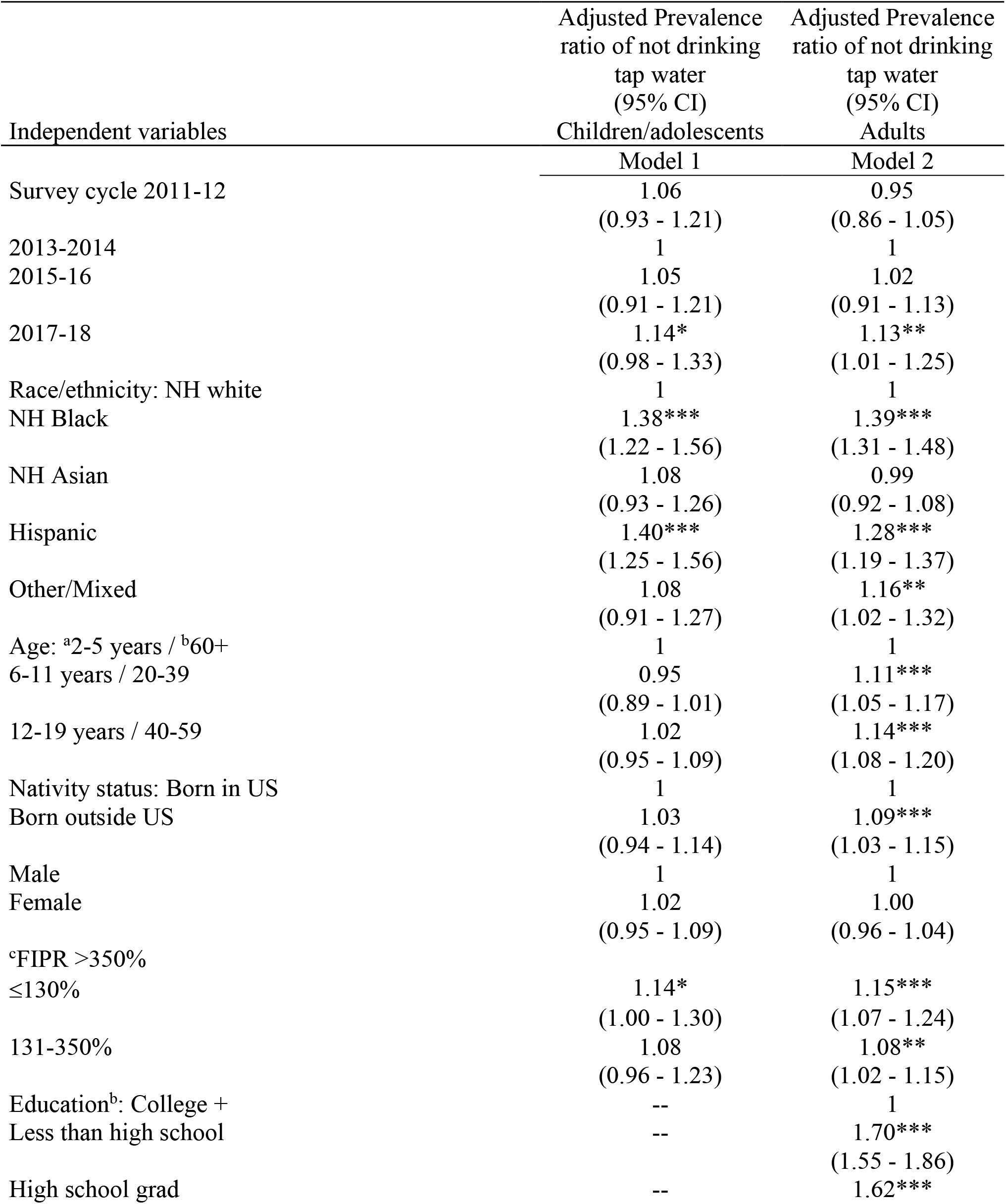
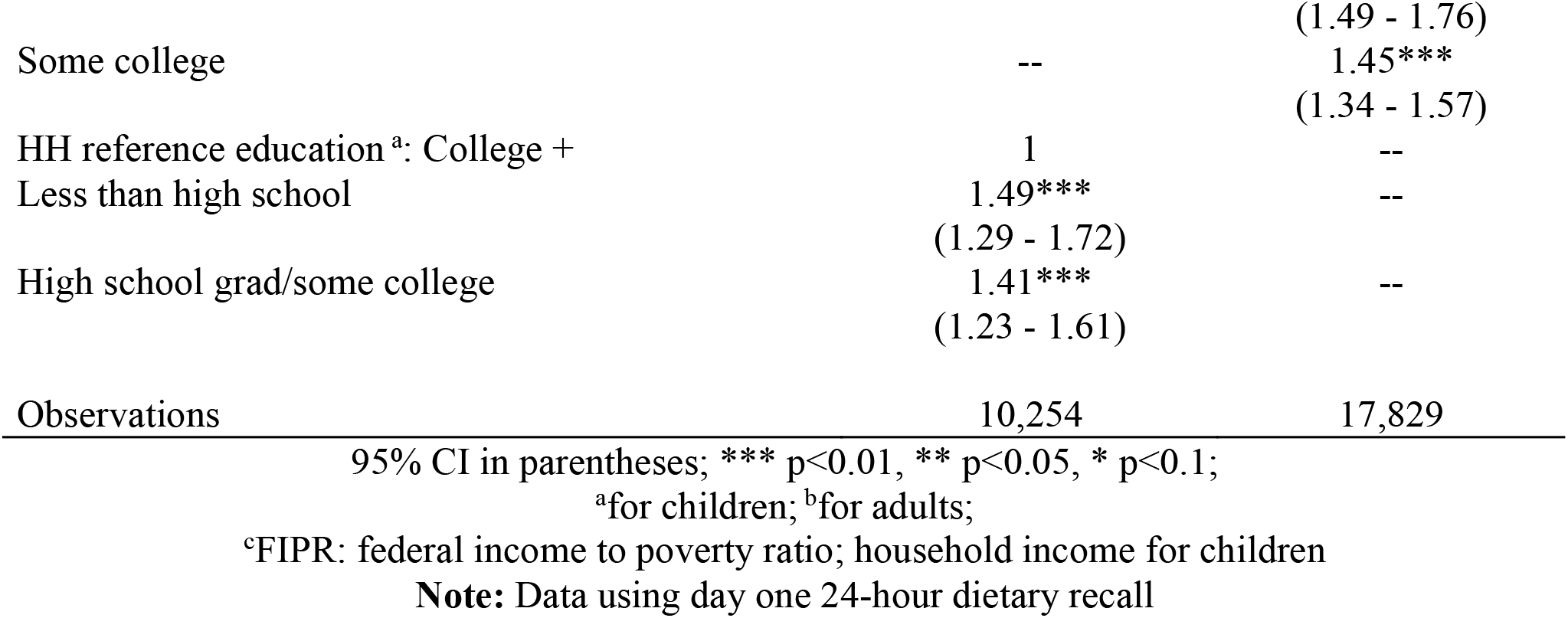
Log-binomial regression models of not drinking any tap water on a given day by survey cycle, race/ethnicity, and socio-demographics, NHANES 2011-2018 among children/adolescents and adults.

**Table 3:**
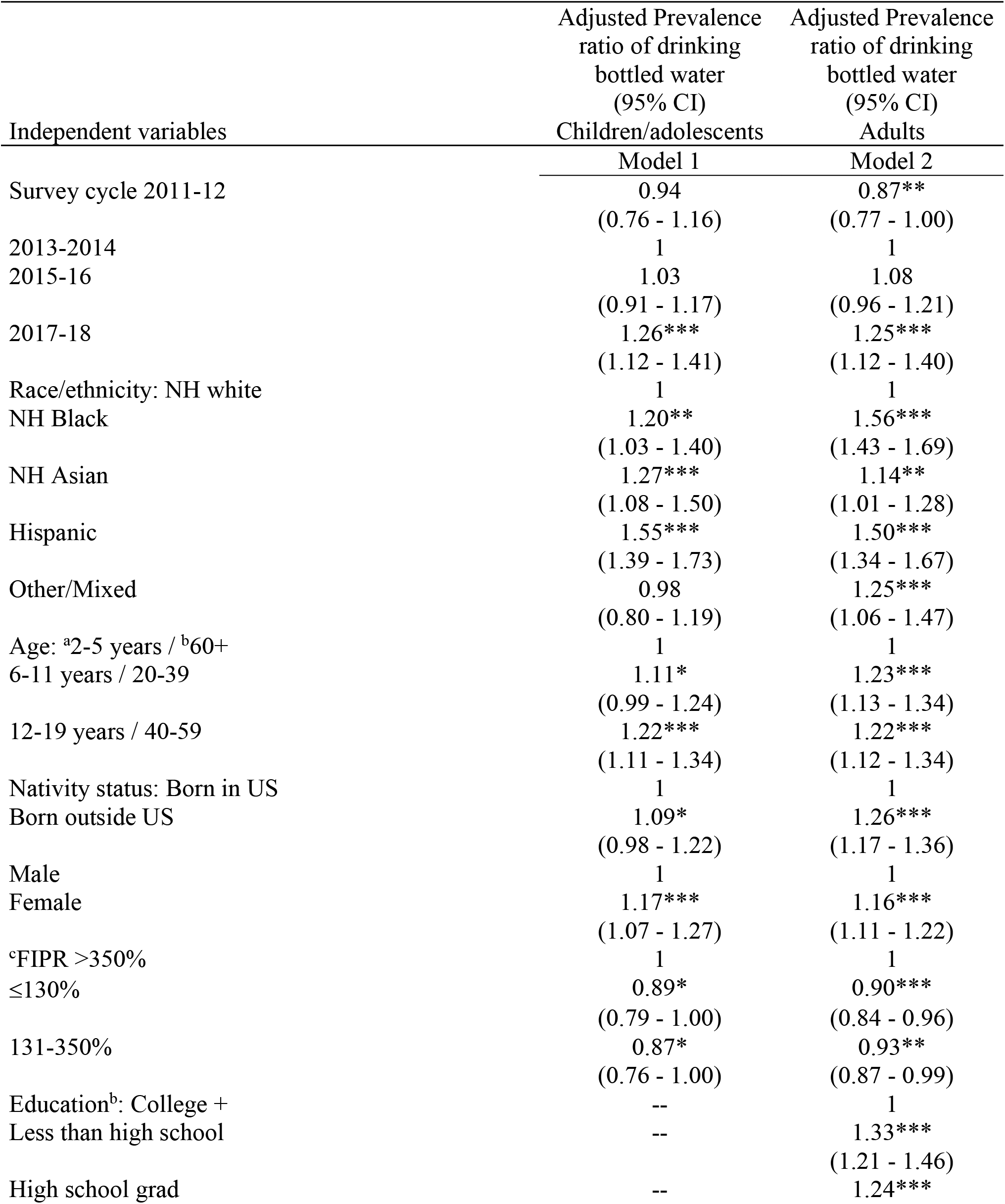
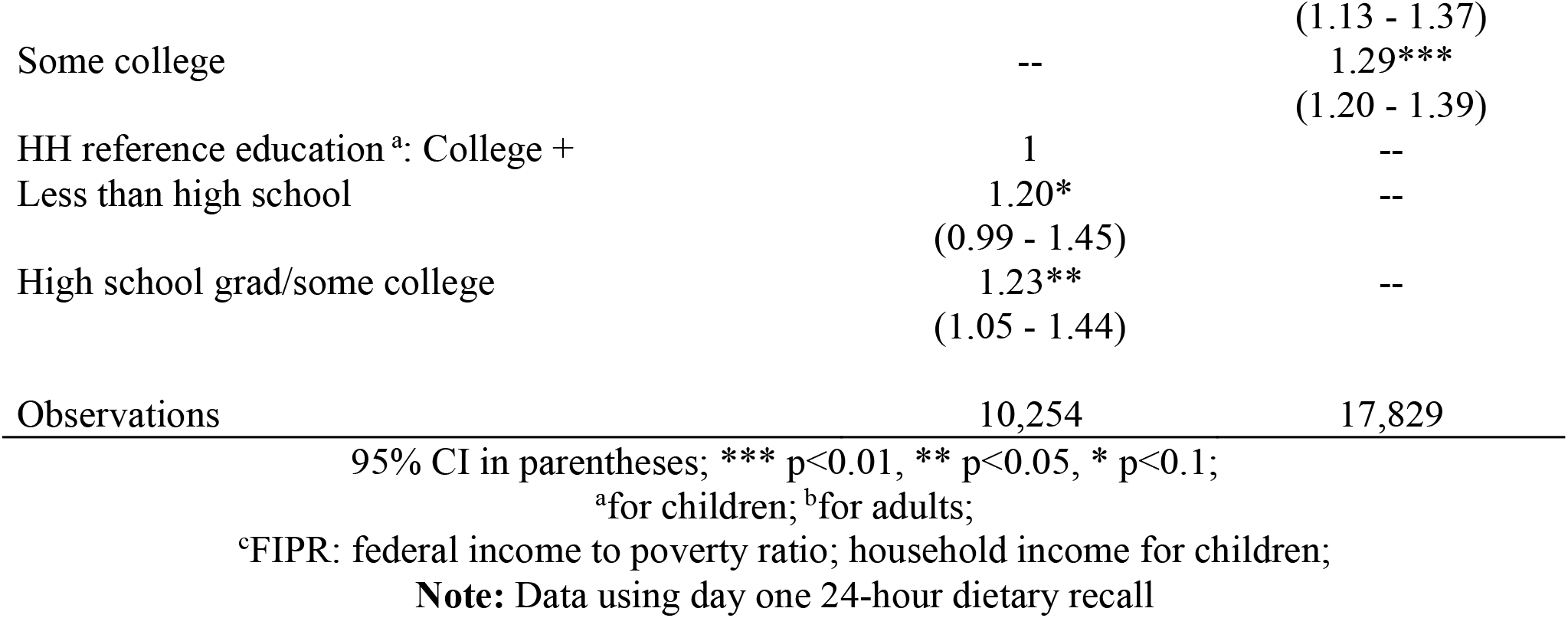
Log-binomial regression models of drinking any bottled water on a given day by survey cycle, race/ethnicity, and socio-demographics, NHANES 2011-2018 among children/adolescents and adults.

In the second analysis, prevalence of any bottled water consumption on a given day was significantly higher overall for both children (PR: 1.26; 95% CI: 1.12-1.41) and adults (PR: 1.25; 95% CI: 1.12-1.40) in 2017-18 than in 2013-14. Further, Black, Asian, and Hispanic persons all had higher prevalence of drinking bottled water than white children and adults, respectively (**Table 4**, Models 1-2).

## Discussion

This study used nationally-representative data to examine trends and racial/ethnic disparities in tap water consumption among US children and adults which included the pre- and post-Flint period as national awareness about water safety concerns increased. There was little change between 2011–14, but probability of not drinking tap water increased successively from 2013-14 to 2017-2018, with Hispanic, Black, and Asian individuals being more likely to not drink tap water than whites. Self-reported race/ethnicity continued to be the largest predictor of not drinking tap water of all covariates examined as the differential treatment of individuals in these categories increases exposure to environmental injustice and racism^11^.

Our findings demonstrate persistent racial/ethnic disparities in the tap water consumption gap and that Hispanic and Black households’ probability of not drinking tap has further increased in recent years. Higher inequities in water access^9^ and distrust^25^ has been pointed to in prior research as reasons that Black and Hispanic populations are less likely to drink tap water^12,19,26^. It is critical for public health nutrition to examine how environmental racism and policies may contribute to health disparities, including disparities in water access and consumption. Tap water consumption has health benefits through fluoride delivery that lowers risk of dental caries^16^. Moreover, when individuals do not drink water they consume double the calories from sugar-sweetened beverages^13^.

Environmental injustice and racism contribute to these disparities^9^ as water quality violations^27^ and water shutoffs^9^ disproportionately affect these communities. Drinking water quality violations often occur at the intersection of race and class where low-income Black and Hispanic communities are at the greatest risk of marginalization^28^. Our results suggest little progress in addressing these inequities. Increased national awareness of water quality violations following the Flint Water Crisis, emerging local water contaminant exceedances in other cities like Newark, NJ, or other unknown factors may help explain our findings. In particular, increasing reports of lead violations in school drinking water, following increasing requirements to test at the tap, have likely heightened concerns regarding tap water safety^3^.

To address these disparities and distrust, expanded water quality testing, communication, and lead remediation efforts are needed. Simplified water quality reports that are easy to understand coupled with clear communication between utility companies and households are paramount (e.g. http://policyinnovation.org/wp-content/uploads/WaterDataPrize_Report.pdf). Further, targeted interventions for marginalized communities that use trusted community stakeholders can help address environmental injustice and rebuild trust^9^.

While recent studies indicate that up to two million Americans lack basic water access^1^ - particularly in urban areas^29^ - our study indicates the broader problem is far more pervasive and growing. In 2017–2018, approximately 20% of US children/adolescents and adults did not drink tap water up from ∼13% in 2013-14. Using the NCHS Census population totals^18^, this translates into 14.8 million children and 46.6 million adults or 61.4 million people in 2017-18 in the US who did not drink their tap water (**Supplemental Table 2**). Since 2013-14, this represents an increase of 19 million people (∼5.6 million children, ∼13.4 million adults) not drinking their tap water, which could be considered an epidemic of tap water distrust and disuse.

Our results differ somewhat from a recent paper examining trends in tap water intake using NHANES 2011-2016 data, which did not find significant changes in the volume of tap water consumed over time, but did uncover increases in the volume of bottled water consumed^30^. While our primary variable was based on whether persons consumed their tap water rather than the average volume of water consumed, our results were robust when using a dichotomous variable of any tap water intake from the 24-hour dietary recall. Second, we found that prevalence of any bottled water consumption on a given day was 25% higher in 2017-18 than 2013-14, supporting the recent trends of increasing bottled water intake in the US^12,30,31^. It is critical for public health nutrition to monitor trends in tap and bottled water use beyond average mean intakes because this can provide a window into potential underlying water insecurity.

Study limitations include the inability to know why participants did not drink their tap water as follow-up questions were not asked. However, we used two measures of tap water consumption to provide additional internal validity of results and find that shifts to bottled water occurred. We were not able to exclude individuals with no access to tap water, however, NHANES samples the civilian, household population^18^ and thus does not sample homeless, who represent a large percentage of those without tap access^32^. This study is limited by not having geographical data, but its goal was to examine recent national trends in tap water consumption rather than geographic differences. Recent work has demonstrated that structural issues like precarious housing within communities of color increases risk of incomplete plumbing^29,33^. The question regarding tap water filtration was discontinued following the 2009-2010 survey and thus was not controlled for. However, prior work has demonstrated that use of tap water filters do not have an overall effect on these national estimates of tap water consumption^12^. NCHS addressed the declining response rates through the sampling weights, which further took into account non-response and loss of screener to examination^18^.

## Conclusion

Approximately 63% and 40% more Black and Hispanic children and adults did not drink their tap water in 2017-18 compared to 2013-14. Overall, that approximately 61.4 million people in the US did not drink tap water, an increase of 19 million since the Flint Water Crisis, represents a substantial opportunity to improve public health nutrition. Further, bottled water consumption continued to increase in the US from 2011–2018, particularly among Black, Asian, and Hispanic households. This reliance on bottled water levies additional socioeconomic strain.

Policies that address the root of environmental injustices that low-income and minority communities face in accessing safe tap water are necessary to halting the growing racial disparity gap observed in tap water consumption^25^. To design meaningful solutions and rebuild trust more work with communities of color is needed to understand the various factors that contribute to eschewing tap water. The field of public health nutrition should engage with utility companies and community stakeholders to design interventions aimed at ensuring access to clean water and reducing these disparities in tap water distrust.

## Data Availability

Data comes from publicly available NHANES and can be downloaded here: https://wwwn.cdc.gov/nchs/nhanes/continuousnhanes/default.aspx?BeginYear=2017

## Supplemental Materials for

**Supplemental Table 1:**
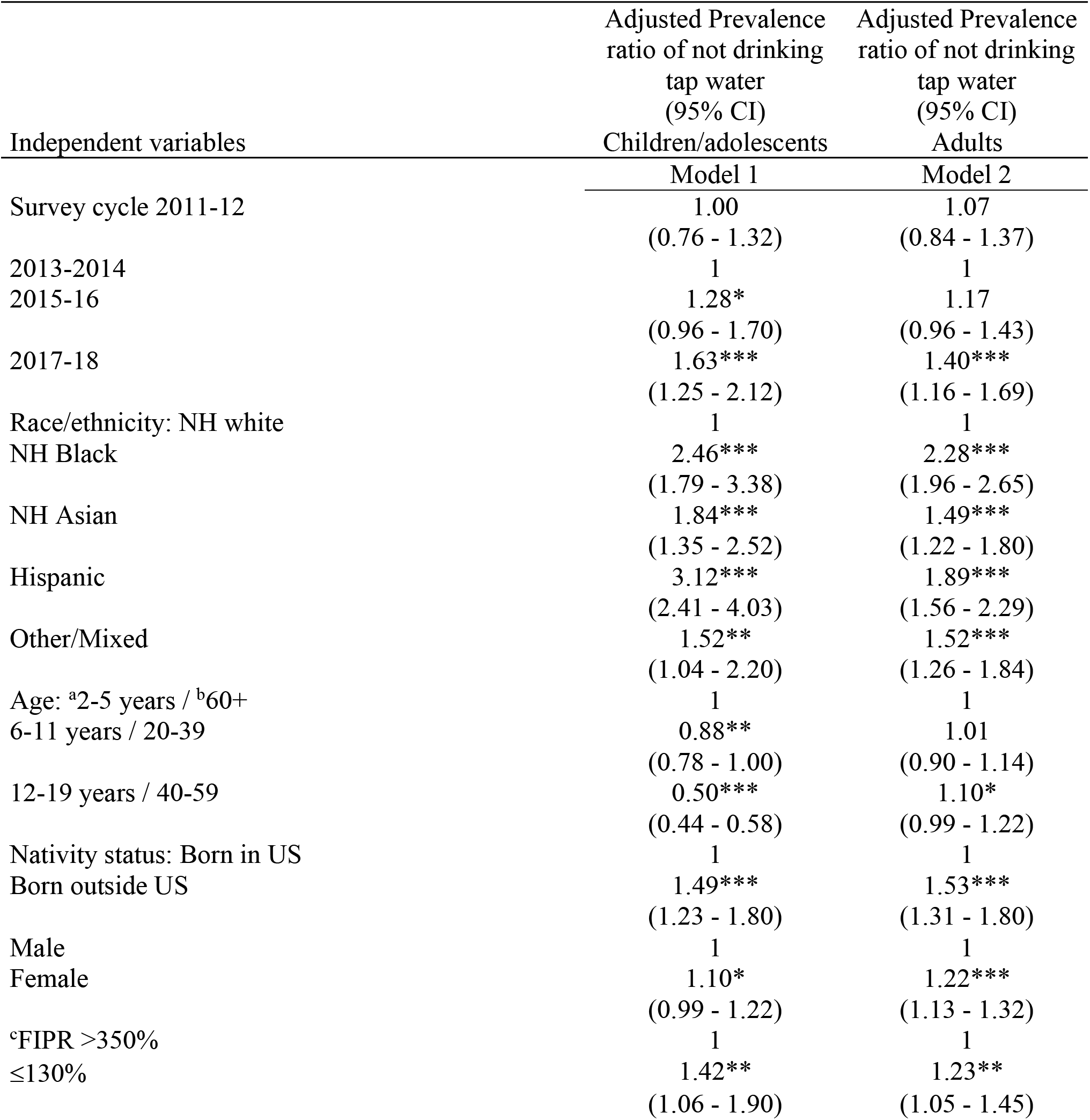
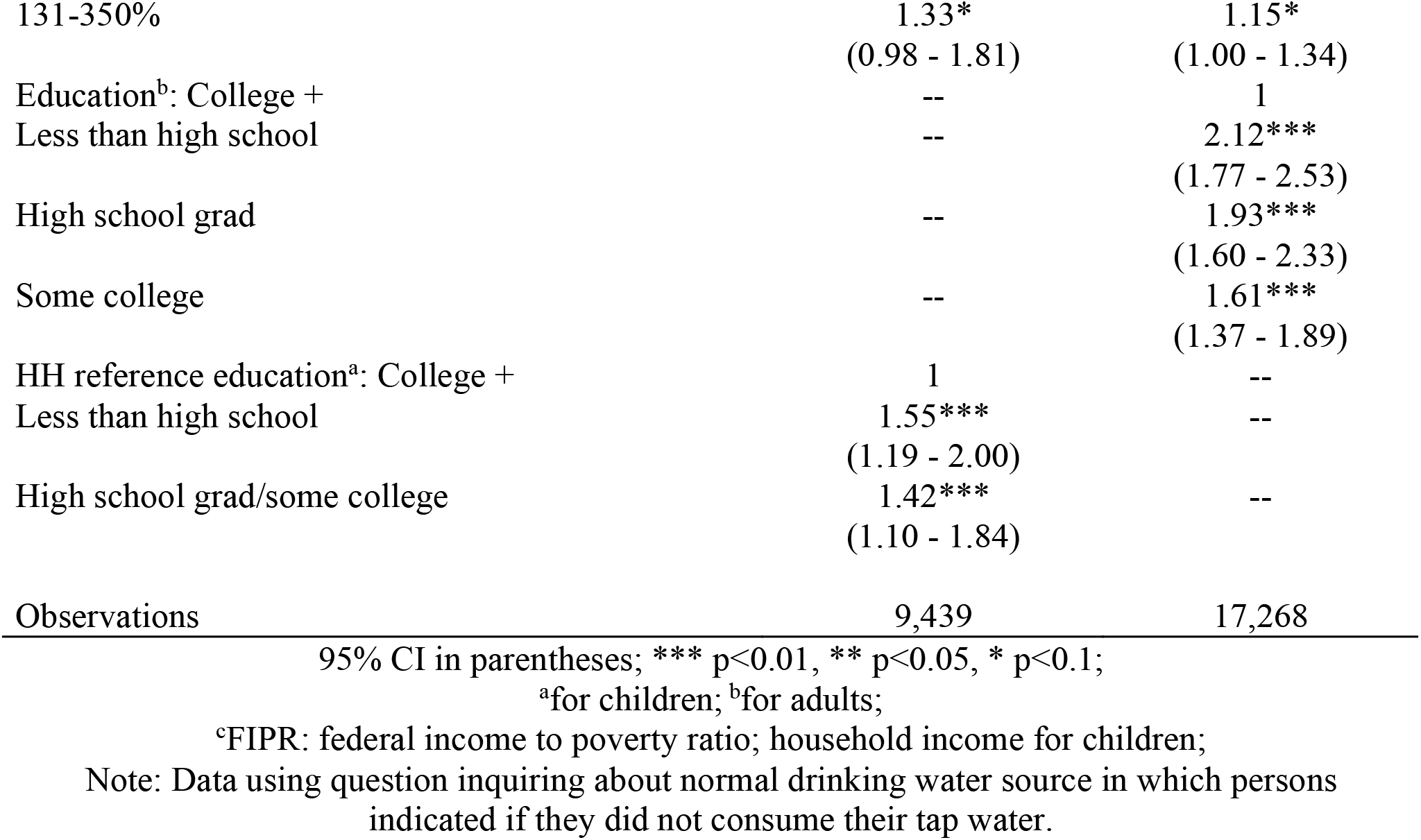
Log-binomial regression models of not drinking tap water by survey cycle, race/ethnicity, and socio-demographics, NHANES 2011-2018 among children/adolescents and adults.

| Independent variables | Adjusted Prevalence<br>ratio of not drinking<br>tap water<br>(95% CI) | Adjusted Prevalence<br>ratio of not drinking<br>tap water<br>(95% CI) |
| --- | --- | --- |
|  | Children/adolescents | Adults |
|  | Model 1 | Model 2 |
| Survey cycle 2011-12 | 1.00<br>(0.76 - 1.32) | 1.07<br>(0.84 - 1.37) |
| 2013-2014 | 1 | 1 |
| 2015-16 | 1.28*<br>(0.96 - 1.70) | 1.17<br>(0.96 - 1.43) |
| 2017-18 | 1.63***<br>(1.25 - 2.12) | 1.40***<br>(1.16 - 1.69) |
| Race/ethnicity: NH white | 1 | 1 |
| NH Black | 2.46***<br>(1.79 - 3.38) | 2.28***<br>(1.96 - 2.65) |
| NH Asian | 1.84***<br>(1.35 - 2.52) | 1.49***<br>(1.22 - 1.80) |
| Hispanic | 3.12***<br>(2.41 - 4.03) | 1.89***<br>(1.56 - 2.29) |
| Other/Mixed | 1.52**<br>(1.04 - 2.20) | 1.52***<br>(1.26 - 1.84) |
| Age: <sup>a</sup>2-5 years / <sup>b</sup>60+ | 1 | 1 |
| 6-11 years / 20-39 | 0.88**<br>(0.78 - 1.00) | 1.01<br>(0.90 - 1.14) |
| 12-19 years / 40-59 | 0.50***<br>(0.44 - 0.58) | 1.10*<br>(0.99 - 1.22) |
| Nativity status: Born in US | 1 | 1 |
| Born outside US | 1.49***<br>(1.23 - 1.80) | 1.53***<br>(1.31 - 1.80) |
| Male | 1 | 1 |
| Female | 1.10*<br>(0.99 - 1.22) | 1.22***<br>(1.13 - 1.32) |
| <sup>c</sup>FIPR >350% | 1 | 1 |
| ≤130% | 1.42**<br>(1.06 - 1.90) | 1.23**<br>(1.05 - 1.45) |
| 131-350% | 1.33* | 1.15* |
|  | (0.98 - 1.81) | (1.00 - 1.34) |
| Education<sup>b</sup>: College + | -- | 1 |
| Less than high school | -- | 2.12*** |
|  |  | (1.77 - 2.53) |
| High school grad | -- | 1.93*** |
|  |  | (1.60 - 2.33) |
| Some college | -- | 1.61*** |
|  |  | (1.37 - 1.89) |
| HH reference education<sup>a</sup>: College + | 1 | -- |
| Less than high school | 1.55*** | -- |
|  | (1.19 - 2.00) |  |
| High school grad/some college | 1.42*** | -- |
|  | (1.10 - 1.84) |  |
| Observations | 9,439 | 17,268 |

95% CI in parentheses; \*\*\* p<0.01, \*\* p<0.05, \* p<0.1;
<sup>a</sup>for children; <sup>b</sup>for adults;
<sup>c</sup>FIPR: federal income to poverty ratio; household income for children;
Note: Data using question inquiring about normal drinking water source in which persons indicated if they did not consume their tap water.

**Supplemental Table 2:**
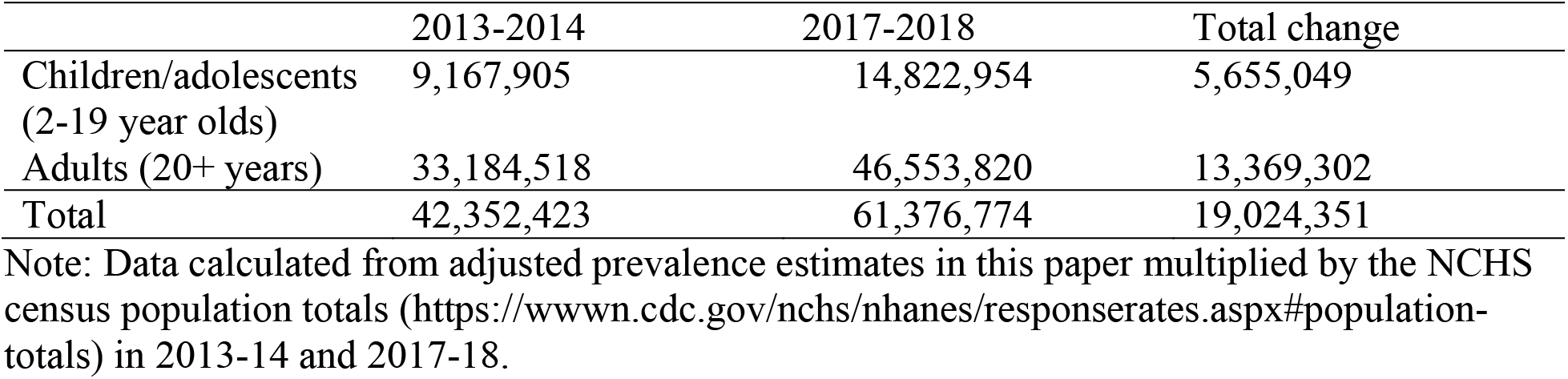
Estimated amount of people in the US not drinking tap water and change between 2013-2014 and 2017-2018.

|  | 2013-2014 | 2017-2018 | Total change |
| --- | --- | --- | --- |
| Children/adolescents (2-19 year olds) | 9,167,905 | 14,822,954 | 5,655,049 |
| Adults (20+ years) | 33,184,518 | 46,553,820 | 13,369,302 |
| Total | 42,352,423 | 61,376,774 | 19,024,351 |

Note: Data calculated from adjusted prevalence estimates in this paper multiplied by the NCHS census population totals (https://wwwn.cdc.gov/nchs/nhanes/responserates.aspx#population-totals) in 2013-14 and 2017-18.

## Notes

**The authors declare they have no conflicts of interest.**

**Funding statement**: This work was supported by the Ann Atherton Hertzler Early Career Professorship funds, and Penn State’s Population Research Institute (NICHD P2CHD041025). The funders had no role in the research or interpretation of results.

### Competing Interest Statement

The authors have declared no competing interest.

### Funding Statement

This work was supported by the Ann Atherton Hertzler Early Career Professorship funds, and the Penn State Population Research Institute (NICHD P2CHD041025). The funders had no role in the research or interpretation of results.

### Author Declarations

NHANES is conducted by the National Center for Health Statistics (NCHS) and approved by their research ethics board. Children aged 7-17 years provided assent and parents provided consent for children under 18; adults consented for themselves.

